# Refinement of post-COVID condition core symptoms, subtypes, determinants, and health impacts: A cohort study integrating real-world data and patient-reported outcomes

**DOI:** 10.1101/2024.06.23.24309348

**Authors:** Yunhe Wang, Marta Alcalde-Herraiz, Kim López Güell, Li Chen, Lourdes Mateu, Chunxiao Li, Raghib Ali, Nicholas Wareham, Roger Paredes, Daniel Prieto-Alhambra, Junqing Xie

**Affiliations:** Nuffield Department of Population Health, University of Oxford, Oxford, UK; Centre for Statistics in Medicine and NIHR Biomedical Research Centre Oxford, NDORMS, University of Oxford, Oxford, UK; Institute of Child and Adolescent Health, School of Public Health, Peking University; Department of Infectious Diseases & irsiCaixa AIDS Research Institute, Hospital Universitari Germans Trias i Pujol, Badalona, Catalonia, Spain; Chair in Infectious Diseases and Immunity, Center for Health and Social Care Research (CEESS), Faculty of Medicine. University of Vic-Central University of Catalonia (UVic-UCC); Universitat Autònoma de Barcelona, Catalonia, Spain; REICOP (Red de Investigación Covid Persistente), Madrid, Spain; Medical Research Council Epidemiology Unit, University of Cambridge, Cambridge, UK; Center for Global Health and Diseases, Department of Pathology, Case Western Reserve University School of Medicine, Cleveland, OH; Department of Medical Informatics, Erasmus University Medical Center, Rotterdam, the Netherlands

## Abstract

**Background:** Post-COVID-19 condition (PCC) affects millions of people, and is an essential component of the long-term impact of COVID-19 during the post-pandemic era. Yet, consensus on clinical case definition and core components of PCC remains lacking, affecting our ability to inform research and evidence-based management. Our study aims 1) to identify the most specific symptoms for PCC , and identify clinical subtypes; 2) to evaluate both virus- and host-related determinants of PCC, and 3) assess the impact of PCC on physical and mental health.

**Methods:** We studied participants from UK Biobank who completed a health and wellbeing survey between June and September 2022. Participants reported the current conditions of the presence, duration, and functional limitations of 45 symptoms, using an online questionnaire designed specifically for COVID-19 research. SARS-CoV-2 infection status and disease history were obtained through linkage to surveillance data and electronic medical records, respectively. Participants reporting symptoms within 30 days after infection were excluded. The most specific PCC symptoms (MSS) were defined using two criteria: statistical significance (P < 0.05 after Bonferroni correction) and clinical relevance (absolute risk increase >5%). Propensity score weighting was used to control for confounding. Subtypes of PCC were then defined based on the MSS among the COVID-19 infected individuals. A multivariable regression was used to study pathogen- and host-related risk factors for PCC, and its impact on 13 physical and 4 mental health patient-reported functional outcomes.

**Findings:** 172,303 participants (mean age 68.9, 57.4% female) were included in the analysis, of whom 43,395 had PCR-confirmed COVID-19. We identified 10 MSS and classified four PCC subtypes: ENT subtype (30.1%), characterized by alterations in smell, taste, and hearing loss; cardiopulmonary subtype (10.4%), characterized by shortness of breath, postural tachycardia, chest tightness, and chest pressure; neurological subtype (23.5%), characterized by brain fog and difficulty speaking; and general fatigue subtype (38.0%), characterized by mild fatigue. A higher PCC risk was observed for patients with Wild-type variant, multiple infections, and severe acute COVID-19 illness, consistently across the four PCC subtypes. In addition, a range of factors, including socioeconomic deprivation, higher BMI, unhealthy lifestyle, and multiple chronic health conditions, were associated with increased PCC risk, except for age and sex. Conversely, vaccination was associated with a largely reduced PCC risk, particularly for the cardiopulmonary subtypes. Individuals with PCC experienced a much worse physical and mental health. Specifically, the cardiopulmonary subtype had the most pronounced adverse impact on function impairments, followed by neurological, mild fatigue, and ENT subtype. The most affected functions included the ability to concentrate, participate in day-to-day work, and emotional vulnerability to health problems.

**Interpretation:** PCC can be categorized into four distinct subtypes based on ten core symptoms. These subtypes appeared to share a majority of pathogen and host-related risk factors, but their impact on health varied markedly by subtype. Our findings could help refine current guidelines for precise PCC diagnosis and progression, enhance the identification of PCC subgroups for targeted research, and inform evidence-based policy making to tackle this new and debilitating condition.

## Introduction

*Post-COVID-19 conditions (PCC)* or *long COVID*, generally defined as ongoing, relapsing, or new onset multisystem symptoms or related conditions during the post-acute stage of SARS-CoV-2 infection, constitute a global public health crisis. This novel clinical entity can persist up to 3 years following the initial infection,^1,2^ which has affected millions of people worldwide,^3^ and had significant impacts on daily functioning, quality of life, work, and healthcare costs.^4^ Unfortunately, limited management strategies have been established to date. There is therefore an urgent need to research PCC-related symptomatology and determinants to inform effective prevention and treatment strategies.

Previous studies with variable designs have reported substantially divergent PCC prevalence and symptoms.^5^ The lack of consensus on case definition and core components of PCC has also led to heterogenous outcome measures,^6^ undermining the comparability between studies and limiting our ability to inform evidence-based management. The four key elements for precision PCC diagnosis should include a core symptom set,^7^ timeframe from infection, persistence, and impacts on daily functioning. However, the adoption of these elements varied significantly across major organizational definitions from WHO, NICE, and the CDC,^8^ and across many previous studies.^5^ In addition, most PCC studies used electronic health records (EHRs) to curate PCC phenotypes, which are known to be limited by heterogeneity in clinical coding, inadequacy and incompleteness of diagnostic codes for the capturing of symptoms, and biases in participant selection and health care use.^9^

PCC should be regarded as an umbrella condition with multiple subtypes, and each may link to distinct physiopathology mechanisms.^10–14^ Yet, previous studies have identified symptom classes that span almost all organ and system domains and were rarely replicable across cohorts. Notably, although PCC has been associated with factors such as age, sex, and ethnicity,^10,11,14^ whether a range of host- and virus-related determinates of overall PCC also consistently related to various phenotypes remains limited. Furthermore, the impact of PCC on patient-reported health and function are still not widely recognised. Accurate refinement of PCC symptomatology, determinants, and health impacts can potentially help reveal the pathophysiology of PCC and inform targeted care strategies.^15^

To fill these knowledge gaps, we used comprehensive data with detailed patient-centred outcomes from participants in a large and rich prospective population-based cohort, and set out to: (1) identify the most specific symptoms (MSS) of PCC and its clinical subtypes; (2) characterize the virus- and host-related determinants of PCC; and (3) assess the impact of PCC on physical and mental health and function.

## Methods

### Data sources

We used data from the UK Biobank, a prospective population-based cohort with over 500,000 participants recruited between 2006 to 2010 across England, Scotland, and Wales. Besides the comprehensive baseline data, several external databases have been linked to UK Biobank participants to enable the follow-up of their health and disease outcomes.^16,17^ Details of the cohort profile and data linkages have been described previously.^18^

During the COVID-19 pandemic, all participants with a valid email address were invited to attend an online Health and Well-being survey. This survey was conducted to assess the impact of COVID-19 by directly collecting data from patients. Initially, a pilot study involving 10,000 participants was first conducted to ascertain the robustness of the survey platform and the acceptability of the questionnaire’s content and length. Subsequently, 333,427 individuals were invited via email, and 60.5% (201,684) completed the online questionnaire between 22^nd^ June 2022 and 15^th^ May 2023. A further 1,094 participants without email invitation completed the survey on the UK Biobank website. The questionnaire included questions on current health issues, mental health conditions, and daily functional impairments, designed by an expert panel with expertise in long COVID. It incorporated items from the Wellcome COVID-19 questionnaire and surveys from other extensive cohort studies such as REACT, TwinsUK, and the ZOE COVID Study. Consultation with the researchers who developed a CORE outcome set for Long COVID was also involved to ensure the inclusion of symptoms most relevant to post-acute COVID. **Supplementary Table 1** lists the origins and sources of questionnaire items.

### Study design

We performed a cohort and a cross-sectional analysis among individuals who had completed the online Health and Well-being survey. For the cohort, we curated an infected group consisting of participants with a prior PCR-confirmed SARS-CoV-2 infection and a control group including individuals without infection, before or at the time of survey administration (*T_s_*). For those with recurrent infections in the infection group, we anchored the most recent infection date as the index date (*T_0_*). To ensure the comparability of follow-up duration (*T_s_*-*T_0_*) between the two groups, individuals in the control group were assigned a random *T_0_* based on the infection group’s calendar time distribution. We excluded people who had an infection within 30 days prior to the survey completion, to ensure the reported symptomatology related to post-acute COVID. Overall, 3% of remaining participants were excluded due to failure of the initial identity check.

For the cross-sectional analysis, we classified long COVID as those who were diagnosed with SARS-CoV-2 and had at least one PCC symptom during the post-acute phase (30 or more days after infection). Two control groups were defined as follows: (1) participants who infected with SARS-CoV-2 yet were asymptomatic during the post-acute phase, and (2) participants with no evidence of SARS-CoV-2 infection before the survey.

### Covariates

We prespecified covariates previously related to COVID-19 for the adjustment of potential confounders based on previous research, including age, sex, index of multiple deprivation (IMD), education, ethnicity, body mass index, smoking status, physical activity level, and all prior comorbidities with prevalence larger than 1% in our cohort.

### Outcomes and measurements

The primary outcomes of the current study were the presence of 45 prespecified somatic and mental symptoms potentially related to COVID-19. This information was collected in the questionnaire based on participants’ responses to the question: “Are you currently suffering from any of the following health issues that are new or have worsened in recent weeks?”. For those reporting symptoms, their impact on daily life and activities was further investigated through the question: “To what extent is the issue affecting you?”, with answer options of ‘It is not limiting me’ and ‘It is causing me to avoid, reduce or spread out my usual activities’. The correlation between these 45 symptoms in 7 domains is provided in **Supplementary Figure 2**.

Secondary outcomes included a predefined list of mental and physical function impairments, identified through the question: “Over the last 2 weeks, how often have you been bothered by any of the following problems?” for mental health using PHQ-4, and “How much difficulty do you have with the following activities?” for physical functioning. Response options were structured on a four-point Likert scale for the former question ranging from ‘Not at all’, ‘Several days’, ‘More than half the days’ and ‘Nearly every day’, and a five-point Likert scale for the later question ranging from ’No difficulty’, ’Mild difficulty’, ’Moderate difficulty’, ’Severe difficulty’, to ’Extreme difficulty’.

We defined answers of ‘Do not know’ or ‘Prefer not to answer’ as missing values. Given that few data were missing, with the highest proportion of missing values observed for the symptoms of hearing loss (2.9%) and bone pain (2.8%), we thus conducted the complete case analysis for this study.

### Statistical analysis

We used propensity score (PS) weighting to control for difference in baseline characteristics between the COVID-19 infected group and the control group. We built a multivariable logistic regression with Lasso L1 penalty to estimate the PS as the probability of belonging to the infection group, using all variables listed above (Covariates). Inverse probability weights were calculated as one divided by the PS in the infection group and divided by one minus PS in the control group. We then examined the covariate balance between groups. An absolute standardized mean difference (SMD) less than 0.1 were regarded as well balanced. The same PS weighting procedures were performed both in the longitudinal and cross-sectional cohort analysis.

We used PS-weighted logistic regression to calculate odds ratio (OR) for the primary and secondary outcomes. We used the most strict Bonferroni method^19^ to correct for multiple testing in our study and reported corrected confidence intervals under the P□<□.05/(45) for the primary outcomes and P□<□.05/(17) for the secondary outcomes. The most specific symptoms (MSS) of PCC were identified using a dual criteria approach^20^: statistical significance (*P* < 0.05 after Bonferroni correction) and clinical relevance (absolute risk increase >5%), after the propensity score adjustment. PCC was defined as the presence of at least one MSS symptom ≥30 days after a COVID-19 infection in the main analysis. PCC subtypes were further classified as the presence of symptoms in each relevant domain, according to existing literature, clinical knowledge, and expert opinion.

We assessed potential determinants for PCC, including pathogen-related factors (SARS-CoV-2 variants, reinfection, hospitalization due to COVID-19) and host-related factors (age, sex, IMD, ethnicity, obesity, lifestyle factor, comorbidity, and COVID-19 vaccination status). SARS-CoV-2 variants were classified according to the calendar period of predominant strain reported in the UK. Lastly, we assessed the impact of PCC on daily functioning based on 13 physical and 4 mental variables.

We conducted two sensitivity analyses. First, we excluded participants who had an infection within 90 (instead of 30) days prior to the survey to define PCC and its subtypes. The adjustment was made as there is no uniform definition for long COVID, which is currently described as conditions occurring 30-90 days after infection in existing guidelines. Second, we defined PCC and its subtypes as the presence of specific symptoms developed 90 days following SARS-CoV-2 infection and last for at least one month with impact on daily functioning, which is largely consistent with the current WHO definition of long COVID.

## Results

### Baseline characteristics of cohorts

Study design and the process of cohort construction are provided in Supplementary Figure 1. Out of 198,768 respondents to the Health and Well-being survey in the UK Biobank, 5,462 individuals were excluded due to either completing the questionnaire after the end date of data linkage (September 30, 2022; 90% of participants completed the survey by August 30, 2022) or having COVID-19 within 30 days prior to the symptom reporting date. Of these, 43,395 were identified with PCR-confirmed COVID-19, constituting the infection cohort, while the remaining 128,908 served as *uninfected controls*. **Supplementary Figure 3** illustrates the distribution of index dates for the infection and control cohorts over the study period (March 2020-Augest 2022). Both groups showed a similar median interval between the index and symptoms reporting dates (215 days in the infection and 218 days in the control). Before weighting, the absolute SMDs for all covariates were below 0.1, except for age: participants in the control group were approximately 2 years older compared with the infection group (mean [SD] age: 69.5 [7.5] years vs 67.3 [7.7] years, SMD). All other socio-demographic factors and comorbidities were similar between groups. Prevalence of chronic diseases were similar between groups. After PS weighting, all characteristics were well balanced (SMD <0.1), and index dates were fully aligned (**Table 1** and **Supplementary Figure 3**).

**Table 1.**
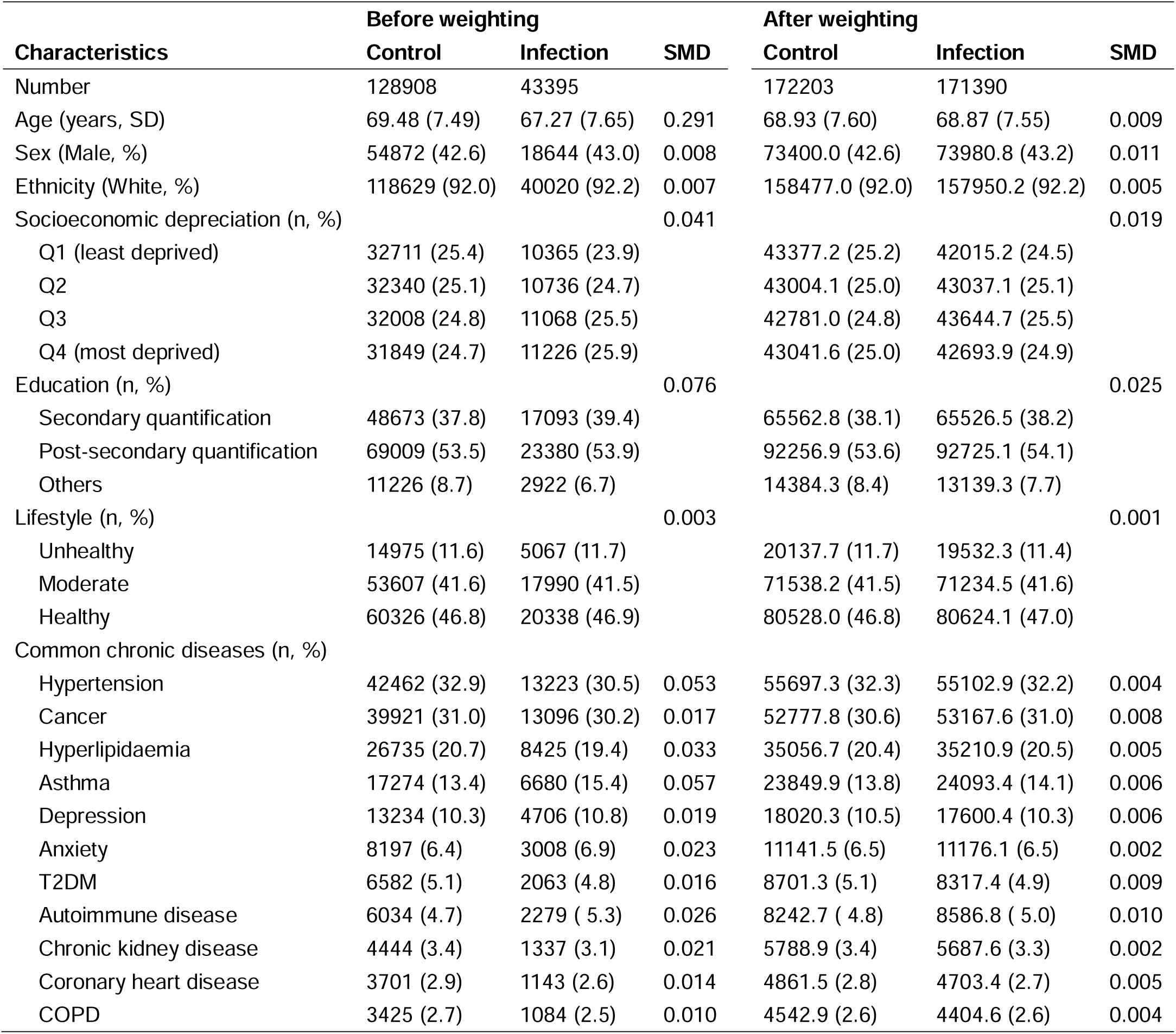
Baseline characteristics of COVID-19 infection vs uninfected control group before and after propensity score weighting.

| Characteristics | Before weighting |  |  | After weighting |  |  |
| --- | --- | --- | --- | --- | --- | --- |
|  | Control | Infection | SMD | Control | Infection | SMD |
| Number | 128908 | 43395 |  | 172203 | 171390 |  |
| Age (years, SD) | 69.48 (7.49) | 67.27 (7.65) | 0.291 | 68.93 (7.60) | 68.87 (7.55) | 0.009 |
| Sex (Male, %) | 54872 (42.6) | 18644 (43.0) | 0.008 | 73400.0 (42.6) | 73980.8 (43.2) | 0.011 |
| Ethnicity (White, %) | 118629 (92.0) | 40020 (92.2) | 0.007 | 158477.0 (92.0) | 157950.2 (92.2) | 0.005 |
| Socioeconomic deprivation (n, %) |  |  | 0.041 |  |  | 0.019 |
| Q1 (least deprived) | 32711 (25.4) | 10365 (23.9) |  | 43377.2 (25.2) | 42015.2 (24.5) |  |
| Q2 | 32340 (25.1) | 10736 (24.7) |  | 43004.1 (25.0) | 43037.1 (25.1) |  |
| Q3 | 32008 (24.8) | 11068 (25.5) |  | 42781.0 (24.8) | 43644.7 (25.5) |  |
| Q4 (most deprived) | 31849 (24.7) | 11226 (25.9) |  | 43041.6 (25.0) | 42693.9 (24.9) |  |
| Education (n, %) |  |  | 0.076 |  |  | 0.025 |
| Secondary quantification | 48673 (37.8) | 17093 (39.4) |  | 65562.8 (38.1) | 65526.5 (38.2) |  |
| Post-secondary quantification | 69009 (53.5) | 23380 (53.9) |  | 92256.9 (53.6) | 92725.1 (54.1) |  |
| Others | 11226 (8.7) | 2922 (6.7) |  | 14384.3 (8.4) | 13139.3 (7.7) |  |
| Lifestyle (n, %) |  |  | 0.003 |  |  | 0.001 |
| Unhealthy | 14975 (11.6) | 5067 (11.7) |  | 20137.7 (11.7) | 19532.3 (11.4) |  |
| Moderate | 53607 (41.6) | 17990 (41.5) |  | 71538.2 (41.5) | 71234.5 (41.6) |  |
| Healthy | 60326 (46.8) | 20338 (46.9) |  | 80528.0 (46.8) | 80624.1 (47.0) |  |
| Common chronic diseases (n, %) |  |  |  |  |  |  |
| Hypertension | 42462 (32.9) | 13223 (30.5) | 0.053 | 55697.3 (32.3) | 55102.9 (32.2) | 0.004 |
| Cancer | 39921 (31.0) | 13096 (30.2) | 0.017 | 52777.8 (30.6) | 53167.6 (31.0) | 0.008 |
| Hyperlipidaemia | 26735 (20.7) | 8425 (19.4) | 0.033 | 35056.7 (20.4) | 35210.9 (20.5) | 0.005 |
| Asthma | 17274 (13.4) | 6680 (15.4) | 0.057 | 23849.9 (13.8) | 24093.4 (14.1) | 0.006 |
| Depression | 13234 (10.3) | 4706 (10.8) | 0.019 | 18020.3 (10.5) | 17600.4 (10.3) | 0.006 |
| Anxiety | 8197 (6.4) | 3008 (6.9) | 0.023 | 11141.5 (6.5) | 11176.1 (6.5) | 0.002 |
| T2DM | 6582 (5.1) | 2063 (4.8) | 0.016 | 8701.3 (5.1) | 8317.4 (4.9) | 0.009 |
| Autoimmune disease | 6034 (4.7) | 2279 (5.3) | 0.026 | 8242.7 (4.8) | 8586.8 (5.0) | 0.010 |
| Chronic kidney disease | 4444 (3.4) | 1337 (3.1) | 0.021 | 5788.9 (3.4) | 5687.6 (3.3) | 0.002 |
| Coronary heart disease | 3701 (2.9) | 1143 (2.6) | 0.014 | 4861.5 (2.8) | 4703.4 (2.7) | 0.005 |
| COPD | 3425 (2.7) | 1084 (2.5) | 0.010 | 4542.9 (2.6) | 4404.6 (2.6) | 0.004 |

### Specific symptoms and PCC subtypes

In PS-weighted cohorts, 17 of 45 symptoms showed positive and statistically significant associations with COVID-19 (BF adjusted P-value <0.05). Among these, seven symptoms did not meet the clinical significance threshold, with an OR <1.05: “tinnitus”, “other hearing issues”, “heart issues”, “joint pain or swelling of joints”, “night sweats”, “unrestful sleep”, and “difficulty sleeping”. The clinically significant ten symptoms, ranked by effect sizes, were: “loss or change in sense of smell”, “loss or change in sense of taste”, “hearing loss”, “shortness of breath or trouble breathing”, “postural tachycardia”, “tightness in the chest”, “chest pressure/heaviness”, “problems thinking (e.g., brain fog)”, “problems communicating (e.g., difficulty speaking)”, and “mild fatigue”. Notably, eight symptoms showed negative and significant associations with COVID-19 (OR<0.95): “nasal congestion”, “sore or painful throat”, “persistent cough”, “phlegm production or chesty cough”, “nausea and/or vomiting”, “decrease in appetite”, “fever (feeling too hot)”, and “chills (feeling too cold)”.

Based on the ten specific symptoms associated with COVID-19, we further ascertained four PCC subtypes: ENT subtype (characterized by alteration in sense of smell and taste and hearing loss), cardiopulmonary subtype (characterized by shortness of breath, postural tachycardia, and chest tightness/pressure), neurological subtype (characterized by brain fog and difficulty speaking), and mild fatigue subtype (characterized by mild fatigue). Overall, 60.1% of participants in the infection group and 57.4% *uninfected controls* had at least one of the ten specific symptoms. The prevalence of PCC subtypes in the infection group was (in order): 38.0% for mild fatigue, 30.1% for ENT, 23.5% for neurological, and 10.4% for cardiopulmonary subtype. These subtypes were also common in the uninfected control group: 36.3% for mild fatigue, 28.0% for ENT, 21.4% for neurological, and 9.52% for cardiopulmonary subtype.

### Pathogen-related factors

The effects of pathogen-related factors with PCC were generally consistent across PCC subtypes (**Figure 2**). The association between COVID-19 and PCC showed great variation across the SARS-CoV-2 variants. The risk for PCC and its subtypes consistently decreased from the initial Wildtype (OR for any PCC 1.32 [95% CI 1.26 to 1.38]) through Alpha and Delta to the most recent Omicron variant (1.07 [1.06 to 1.09]). The only exception was for the ENT subtype, where the Delta variant showed the highest risk (Delta 1.36 [1.32 to 1.41] vs Wildtype 1.27 [1.21 to 1.33]). Participants with multiple infections (1.41 [1.33 to 1.50]) had a higher risk of any PCC compared to those with a single infection (1.10 [1.09 to 1.12]). Similarly, those hospitalized due to COVID-19 within 30 days post-infection (OR 2.01 [95% 1.09 to 2.12]) showed a consistently increased risk of PCC compared to non-hospitalized participants (1.09 [1.07 to 1.11]).

**Figure 1.**
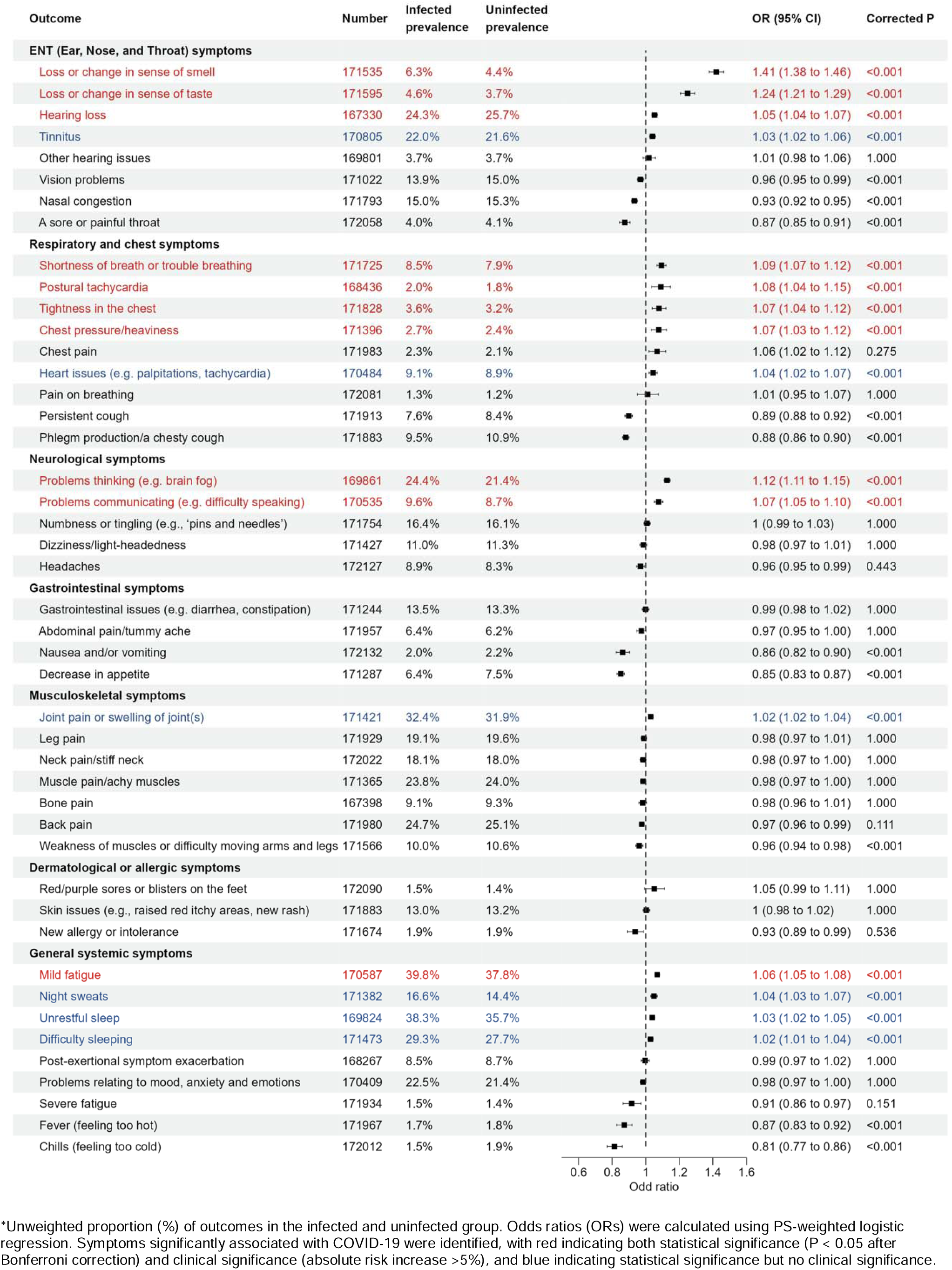
Associations of COVID-19 with the presence of 45 symptoms potentially related to PCC during the post-acute phase of infection.

**Figure 2.**
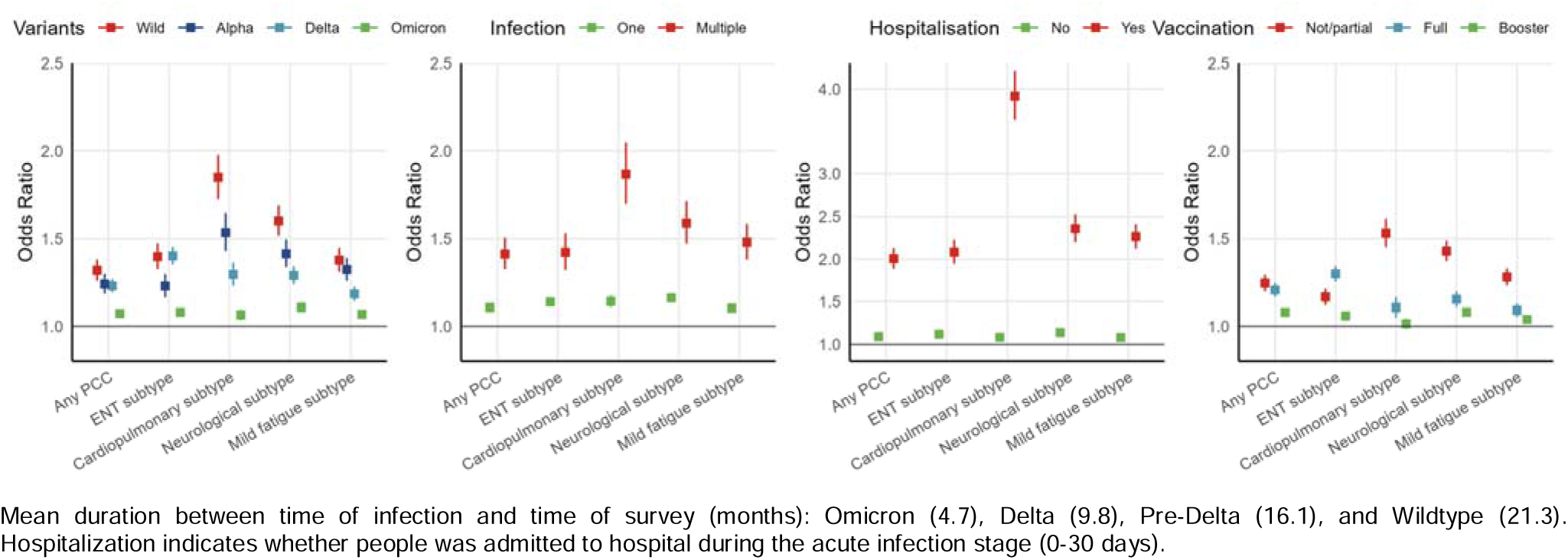
Associations of SARS-CoV-2 pathogen-related factors with PCC and its subtypes.

These associations were consistently observed across PCC subtypes. Full vaccination and booster were associated with significantly lower risk of PCC compared to no or partial vaccination (no/partial 1.25 [1.20 to 1.29] vs booster 1.08 [1.06 to 1.10]). Risk of ENT subtype was higher in participants with full vaccination, coinciding with the dominance of the Delta variant during the period of increasing vaccine uptake (data not shown). Notably, wildtype infections, multiple infections, and hospitalization due to COVID-19 were more strongly associated with the cardiopulmonary PCC subtype, suggesting that the number and severity of acute SARS-CoV-2 infections may be more directly and biologically related to post-acute cardiopulmonary symptoms than multiple extrapulmonary sequelae.

### Host-related factors

A range of sociodemographic and clinical factors were significantly associated with the risk of PCC, with similar effects observed across PCC subtypes for most factors except for age and sex (**Figure 3**).

**Figure 3.**
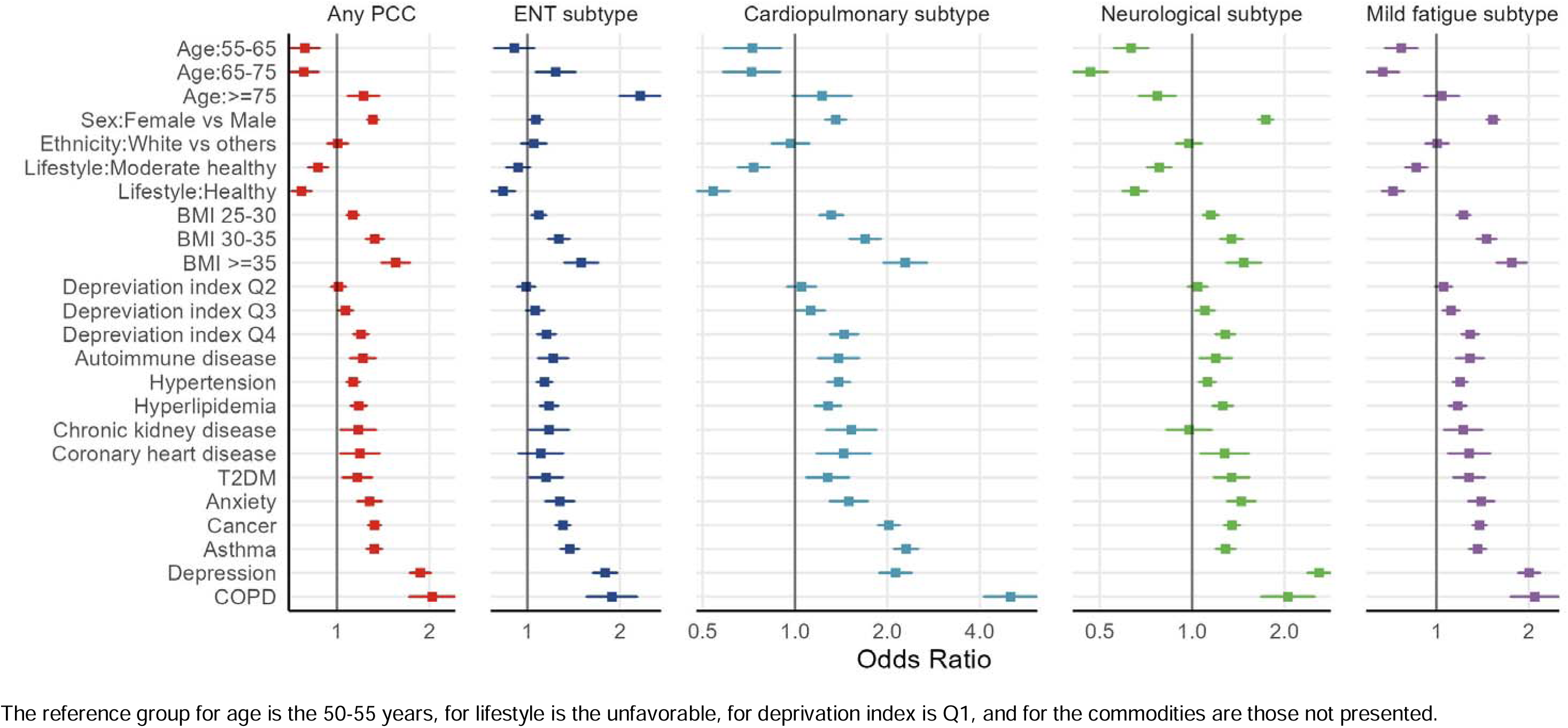
Associations of host-related factors with PCC and its subtypes.

Compared with those aged 45-55 years, participants aged 55-65 (OR 0.79 [0.71 to 0.88]) and 65-75 (0.79 [0.70 to 0.88]) years were at a lower risk of any PCC, while those over 75 years were at increased risk (1.23 [1.09 to 1.39]). However, the age patterns varied across PCC subtypes. The risk of ENT subtype increased with a gradient of increasing age, whereas participants with neurological and mild fatigue subtypes were more likely to be younger. Females were at increased risk for PCC overall (OR 1.29 [1.24 to 1.35]), and for cardiopulmonary, neurological, and mild fatigue subtypes, but at lower risk of ENT subtype (0.88 [0.84 to 0.93]). Overall, PCC risk increased with socioeconomic deprivation, with an OR of 1.20 [1.14 to 1.28] for those living in the most deprived areas. No difference in risk of PCC were seen across ethnic groups, except for the ENT subtype.

The risk for PCC overall and across subtypes increased monotonically with BMI, with OR ranging from 1.50 (95% CI 1.32 to 1.71) for ENT to 2.29 (1.94 to 2.70) for cardiopulmonary subtypes in those with BMI ≥35 compared to BMI <25. Adherence to a healthy lifestyle was consistently associated with lower risk of PCC overall (OR 0.63 [0.58 to 0.67]) and subtypes. Vaccination was associated with a significant reduction in the risk of PCC, overall and by subtype.

All studied comorbidities showed a positive association with PCC (overall and subtypes), except for chronic kidney disease in relation to neurological PCC (OR 0.99 [0.83 to 1.17]). COPD and depression appeared to be the most significant comorbidities related to the risk of PCC (OR 2.05 [1.72 to 2.44] and 1.89 [1.75 to 2.04], respectively), with COPD having the strongest effect on cardiopulmonary subtype (5.04 [4.12 to 6.16]) and depression on neurological subtype (2.24 [2.08 to 2.40]). The prevalence of comorbidities by PCC status is provided in **Supplementary Figure 4**.

### Impact on physical and mental health and function

Individuals with PCC experienced a marked impact on all measured physical health/function metrics, although the magnitude of associations varied considerably across PCC subtypes and outcomes (**Figure 4a**). The ENT subtype was generally associated with the smallest impact on physical function (highest OR: 6.42 [5.97 to 6.90] for “doing day to day work”), followed by mild fatigue subtype (highest OR 9.35 [8.48 to 10.31] for “doing day to day work”), and cardiopulmonary subtype (highest OR 14.71 [13.64 to 15.87] for “doing day to day work”). Notably, the neurological subtype had the strongest negative impact (highest OR: 16.58 [15.11 to 18.19] for “concentrating on doing something for ten minutes” among 13 difficulties).

**Figure 4.**
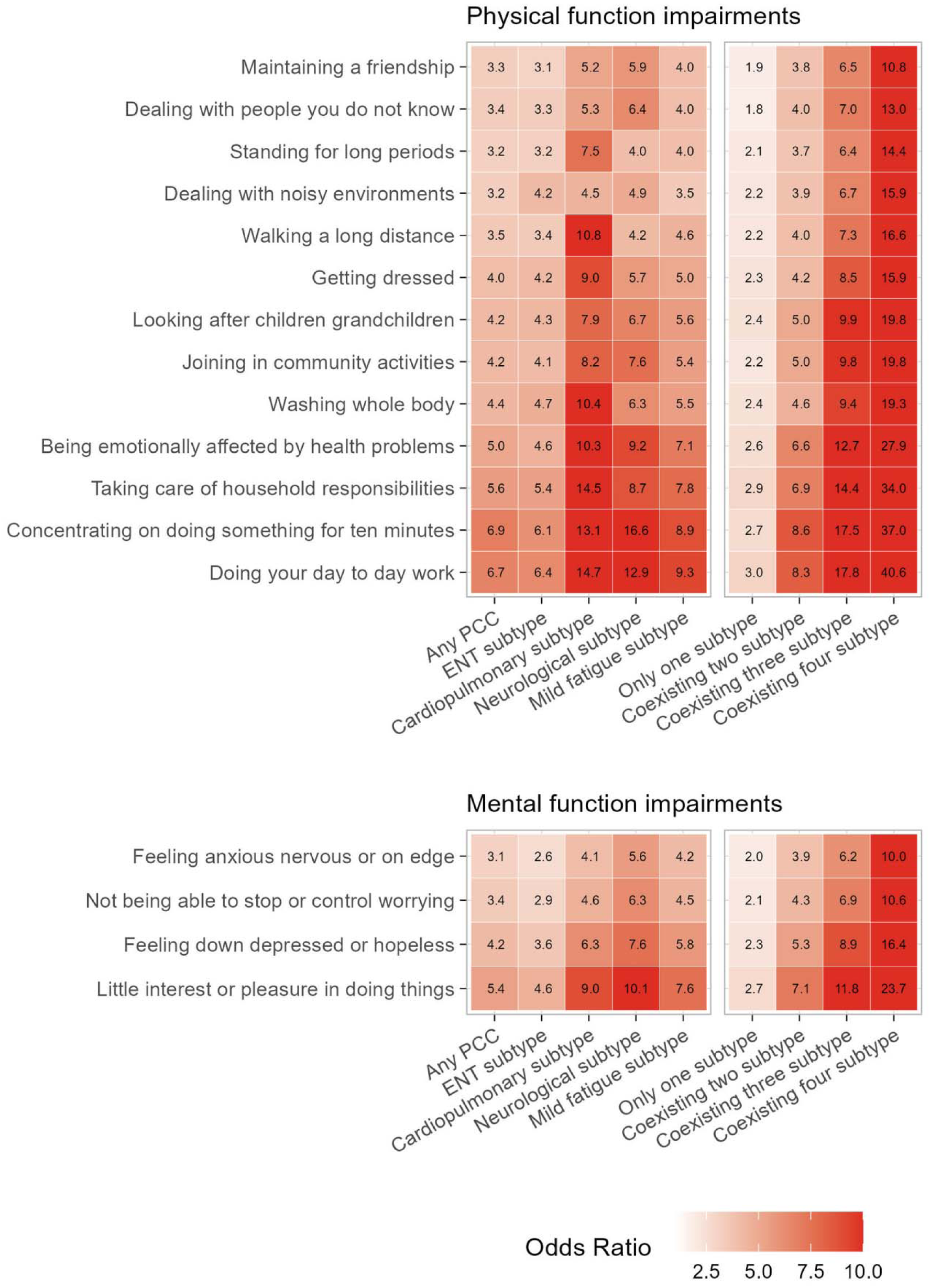
Associations of PCC and its subtypes with physical and mental function impairments.

Across these physical outcomes, the “being emotionally affected by health problems”, “taking care of household responsibilities”, “doing day to day work”, and “concentrating on doing something for ten minutes” were the four physical functions most adversely affected by PCC and its subtypes.

People with PCC also experienced a great impact on mental health and function/s (**Figure 5b**). Although the associations with mental health appeared to be less pronounced than with physical functions, the patterns of association remained similar across PCC subtypes. For instance, the ENT subtype generally showed the lowest OR across mental functioning outcomes, with the neurological subtype had the strongest impacts.

Generally, a higher number of coexisting PCC subtypes led to worse impacts on physical and mental health. Adjusted ORs for physical outcomes ranged from 1.83 [1.70 to 1.96] to 3.04 [2.82 to 3.27] in those with two concurrent PCC subtypes, and from 10.78 [9.77 to 11.90] to 40.58 [36.80 to 44.76] in those with four concomitant subtypes. Similarly, OR of mental outcomes ranged from 1.96 [1.88 to 2.05] to 2.70 [2.53 to 2.88] in those coexisting with two subtypes, and from 10.59 [9.77 to 11.48] to 23.73 [21.69 to 25.95] in those with four subtypes.

Sensitivity analyses using definitions based on 90-day windows identified broadly similar symptomatology patterns (**Supplementary Figure 5**). Prevalence of 45 PCC-related symptoms in participants with and without COVID-19 by definition are provided in **Supplementary Figure 6**.

## Discussion

In this comprehensive study integrating both longitudinal electronic medical records and patient-centred data among a large community-based cohort, we discerned the ten most indicative symptoms for the diagnosis of PCC. Based on the identified symptomatology, we classified PCC into four major subtypes, dominated by ENT, respiratory and cardiac symptoms, cognitive impairments, and mild fatigue. In addition, we provided an in-depth evaluation of the determinants of each PCC subtype, including both virus- and host-related aspects: infection by pre-omicron variants, repeat infection, severe acute COVID-19, overweight and obesity, unhealthy lifestyle, and multiple comorbidities significantly increased PCC risk across all subtypes. Conversely, vaccination against COVID-19 (primary and booster) significantly reduced the risk of all PCC subtypes.

Pre-existing comorbidities were more closely associated with PCC symptoms affecting the same organs/systems. For example, COPD and depression led to higher risks of cardiopulmonary and neurological subtypes, respectively. Lastly, our results reveal that the cardiopulmonary and neurological subtype had the strongest adverse impacts on both physical and mental health. These results highlight the heterogeneity in the presentation of PCC, demographic and clinical risk factors, and related functional impact of PCC, and the importance of characterizing the underlying mechanisms. These novel findings have great potential to advance the current understanding, diagnosis, and management of the long-term adverse effects of COVID-19 during the post-pandemic era.

### Main findings in context

#### Symptomatology of PCC

Despite notable difference in target populations and methodologies used (data source, study design, follow-up length, and analytical approach), the symptomatology patterns across four major PCC subtypes identified in the current study are highly aligned with prior key studies. For example, a cohort study in the Netherlands utilised repetitive cross-sectional design. It proposed 10 core PCC somatic symptoms,^21^ including respiratory-cardiac symptoms, olfactory-auditory symptoms, and general fatigue. Another similar longitudinal study in Switzerland identified 6 symptoms that were at the most excess risk 6 months after infection (“altered taste or smell”, “dyspnoea”, “post-exertional malaise”, “reduced concentration”, “memory”, and “fatigue”).^22^ Also, a population-based study of German participants found four symptom clusters, including fatigue, neurocognitive impairment, chest symptoms, and smell or taste disorders.^23^ Similarly, a UK-based study found the top 7 symptoms at 12 weeks following infection were loss or change of sense of smell or taste, shortness of breath, fatigue, difficulty thinking or concentrating, chest tightness or pain, and poor memory.^24^ Together, these consistent findings promise the utility of the identified MSS for the accurate assessment of patients with PCC in routine clinical practice, and for the effective recruitment of trial participants in future research.

Differences, however, were also noted. Many studies have associated gastrointestinal symptoms, such as dyspepsia and constipation, and musculoskeletal symptoms, such as joint, muscle or back pains, with COVID-19 following the acute infection.^14,25,26^ Most of these studies were conducted based on routinely collected data from electronic health records (EHRs). There is a growing awareness that capturing the symptomatology through real-world clinical data could be challenging.^9^ Also, due to the nonspecific nature of symptoms, most are often not well-represented by diagnostic coding schemes such as ICD-10. Our study analysed comprehensive self-reported data and found neither one of the gastrointestinal or musculoskeletal symptoms to be significantly associated with PCC, although they tended to be very common after COVID-19 and in those without infection. Feeling hot/cold, an acute COVID-19 symptom, was also suggested as a core PCC symptom,^21^ but it was negatively associated with PCC in our study. Interestingly, the negative association was consistently observed for other typical acute symptoms, such as sore throat, cough, nausea, and appetite decrease. This phenomenon might be explained by individual’s perception change over time since the infection: infected people could be overly attentive to typical acute symptoms during the early phase of infection, which may in turn result in a significant reduction of reporting the same symptoms once physically recovered or psychologically relieved.^27^ More investigations are needed to clarify this potential self-reporting bias in research focusing on PCC symptoms.

#### Host- and virus-related determinants

Our study also provides novel insights into the determinants for PCC development. Across PCC subtypes, the effect pattern of virus-related factors appears consistent, yet often varies by host-related factors. The effects of most risk factors across subtypes, such as deprivation, unhealthy lifestyle, obesity, and comorbidities, are consistent with previous findings that mainly do not distinguish PCC subtypes, suggesting the health inequalities in PCC and the potential benefits of adherence to a healthy lifestyle in its prevention. However, heterogenous effects were observed for age and sex. A meta-analysis of 41 studies suggested that older age and female sex are independent risk factors for PCC.^28^ Our findings generally support these associations but revealed that their effects may differ depending on the specific PCC subtype, with younger age related to higher risk of neurological and fatigue subtypes. This variation may partially explain the prior inconsistent evidence in this regard, where data from the UK Office for National Statistics showed that the middle-aged adults have the highest prevalence of PCC,^29^ suggesting that aging might have a two-way effect. Similarly, while females had a higher PCC risk than males, this may not be uniformly applicable to all PCC subtypes, as ENT subtype appears more prevalent in men. Additionally, our findings into specific comorbidities and risk for different PCC subtypes has implications for the management of PCC. Organ-specific comorbidities such as COPD and depression were more closely related to corresponding cardiopulmonary and neurological subtypes, highlighting that baseline organ-level deficits can affect PCC subtype/s risk.

Virus-related factors have relatively consistent effects on PCC and across subtypes. The risk of PCC following infections of different SARS-CoV-2 variant is not yet conclusive, with comparable or lower PCC risk following Omicron infection compared with Delta and other variants.^30,31^ Our study robustly suggested that individuals who first infected during the Omicron-dominated period are at the lowest PCC risk across subtypes, despite the shortest time window since the infection. Nevertheless, the PCC risk remains in individuals following Omicron infection as compared to those uninfected, it is thus foreseeable that the absolute number of new PCC cases would likely to keep accumulating as infections continue. Moreover, our study confirmed further that reinfection and disease severity as important risk factors for PCC^32,33^ and subtypes, especially the cardiopulmonary sequelae. Importantly, people infected after receiving the primary two-dose vaccination were at a significantly lower risk than those who were unvaccinated or only partially vaccinated. However, their risk remained relatively higher than those who had received booster doses. Together, this finding provides further evidence that vaccination may offer protection beyond the acute illness to post-acute complications.^34^ Given that the SARS-CoV-2 variants and vaccination status are two highly correlated conditions, it is hardly to disentangle their individual effects in our study but warrants further research.

#### Physical and mental function impairments

Evidence on the functional impact of PCC was primarily based on survey studies, with high prevalence of functional limitations reported among individuals who self-reported living with PCC.^4^ For example, 78.5% of participants from the US Household Pulse Survey, 58.6% from the UK ONS, and 21.3% from the Canadian national survey experienced daily activity limitations.^4^ However, the lack of a matched control group limited our ability to infer the direct impact of PCC, given that most of these outcomes were also common following COVID-19 and in the general population during the pandemic. Other studies with control groups consisted of participants mainly from post-COVID-19 support groups or used self-diagnosed PCC, undermining the generalizability and validity of findings due to selection and self-reporting biases. After accounting for a comprehensive list of underlying comorbidities, behavioural and sociodemographic factors, we found that PCC and its subtypes were associated with a range of physical and mental health impairments. Overall, the most commonly affected functions were the ability to concentrate, participate in day-to-day work, and take care of household; participants with PCC were also more emotionally vulnerable to health problems and had worse general mental health. Our data suggested that neurological PCC had the most significant adverse impact on physical and mental functions. Although the increased risk of a range of multiorgan symptoms was reported during the 6 to 12 months following SARS-CoV-2 infection,^1^ only the increased risk of cognitive deficit (brain fog) and dementia persisted up to two years.^35^ These findings suggest that service provision and mitigation strategies on the neurological sequelae of COVID-19 should be prioritised and sustained. Severity and subtypes of PCC should be considered when providing targeted care and disability support services.

#### PCC subtypes with varying symptomatology, risk factors, and function impairments

The four major PCC phenotypes identified based on patient-centred data, including the ENT, cardiopulmonary, neurological and general fatigue subtypes, as well as their impairments on physical and mental function, corroborate the current empirically-based consensus for the core outcome set (COS) of PCC.^7^ In the Delphi consensus study by both investigators and patients with lived experience of PCC, the COS included “functioning, symptoms, and conditions” for the cardiovascular, respiratory, nervous system, cognitive, mental health outcomes, and fatigue and pain in the physiological or clinical outcomes domain, and two physical outcomes in the life impact outcomes including work or occupational and study changes and other changes in physical function.^7^ Our study showed consistent symptomatology patterns and function impairments across multiple domains, with the exception of pain. It is likely that the preferred outcomes by patients with lived experience might differ from outcomes considered important by investigators or clinicians,^7^ especially considering that non-specific pain is one of the most common symptoms of PCC.^5^ We confirmed an increased risk of joint pain following post-acute infection, though with low clinical relevance.

Further, the phenotypes identified in our study using patient-reported symptom data largely align with symptom clusters in two nationally representative studies.^13,14^ A study of non-hospitalized adults in England^14^ identified three clusters from 50 consolidated symptoms using primary care medical records: class 1 dominated by a range of non-specific symptoms including pain, fatigue and rash (80.0%), class 2 dominated by respiratory symptoms such as cough and shortness of breath (5.8%), and class 3 dominated by neurological and mental health outcomes such as depression, anxiety, insomnia, and brain fog (14.2%). An US study using RECOVER cohort^13^ also found that four clusters featured by olfactory dysfunction, cardiopulmonary sequelae, neurocognitive impairment, and fatigue. Similarly, these findings do not support the musculoskeletal and digestive symptom clusters previously prioritised in most EHR-based studies,^11,12^ although similar cardiopulmonary and neurological subtypes were also identified in these studies.

The effects of socioeconomic and clinical risk factors were largely consistent across the symptom clusters, however, varying effects of demographics such as age, sex, and ethnicity were observed in the current and previous studies.^10,11,14^ Distinct gene expression patterns during acute infection and inflammation signatures were linked to different symptom clusters, highlighting the heterogeneous etiologies of PCC subtypes.^36,37^ In light of our findings and previous evidence, heterogenous phenotypes of PCC with varying distinct patterns of symptoms, risk factors, and related burden of function impairments need to be considered in the definitions, diagnosis, and management of PCC; there is also the need to better understand underlying mechanisms linked to various phenotypic subtypes and develop targeted therapies for different subtypes using precision medicine to improve outcomes for people living with PCC.

### Strengths and limitations

The major strengths of this study are the size and prospective nature of our community-based cohort, and the richness of information available on longitudinal health, comorbidities, and on self-reported symptoms. The bespoke study design not only allowed for a prospective comparison between COVID-19 patients and concomitant uninfected controls enrolled from the same source population, but also a thorough adjustment for multiple covariates. This thus enabled us to minimise confounding, an issue frequently seen in prior EHR-based studies on this topic. Moreover, all the assessed PCC symptoms were pre-determined based on a consensus from an expert panel, and the resulted questionnaire has been validated. Lastly, the integration of multi-source clinical data with patient-reported outcomes enables the research of different risk factors and consequences of PCC within a large-scale cohort. These methodological features collectively strengthen the validity of our study findings.

Our study findings should be interpreted with caution in the context of some limitations. First, UK Biobank participants are likely to be older and healthier than the UK general population, and are mostly White European, which may limit the generalizability of study findings. Despite the relative risk between comparison groups was largely not influenced, the absolute estimate of symptomatology prevalence should be interpreted with caution. Second, despite we used a robust statistical approach and a range of covariates to account for potential differences in characteristics between groups, residual confounding and reverse causality cannot be totally ruled out in this observational study. Third, a proportion of participants in the uninfected control group may have undiagnosed or untested COVID-19. However, this tends to underestimate the effect size and leads to more conservative results of symptom selection. Fourth, the selection of symptoms most specific to long COVID from a prior consensus symptom list was driven by both statistical significance and clinical relevance. However, it is likely that several unselected symptoms such as general pain may also be more commonly reported in patients with PCC if the baseline risk factors such as comorbidities influence the likelihood of healthcare use and symptom reporting. Further guidelines are needed to integrate evidence from patients, investigators, and clinicians for better management of PCC phenotypes and their impacts.

## Conclusions

We identified four PCC subtypes with distinct symptomatology, functional impact, and some differential risk factors. PCC affected mental and physical health and functions significantly. Our findings will guide future assessment, management, and research on PCC as we aim to better understand distinct mechanisms linked to PCC; and ultimately to develop targeted therapies to improve the patient outcomes.

## Data Availability

We used data from the UK Biobank, a prospective population-based cohort with over 500,000 participants recruited between 2006 to 2010 across England, Scotland, and Wales. Besides the comprehensive baseline data, several external databases have been linked to UK Biobank participants to enable the follow-up of their health and disease outcomes.16,17 Details of the cohort profile and data linkages have been described previously.18

